# Are we in a Big Data era for multiple sclerosis? Lessons from integrating clinical trials and observational studies data into the PRIMUS precision medicine platform

**DOI:** 10.1101/2024.10.17.24315655

**Authors:** Stanislas Demuth, Igor Faddeenkov, Julien Paris, Olivia Rousseau, Béatrice Baciotti, Marianne Payet, Romain Casey, Sandra Vukusic, Senan Doyle, Guillaume Jarre, Nicolas Vince, Sophie Limou, Jérôme De Sèze, Anne Kerbrat, David Laplaud, Gilles Edan, Pierre-Antoine Gourraud, the PRIMUS Consortium

## Abstract

**Objective:** The “Projections In Multiple Sclerosis” (PRIMUS) project aims to develop a precision medicine platform enabling neurologists to support therapeutic decisions in multiple sclerosis by visualizing similar patient data among a reference database. We present a data integration method to combine randomized clinical trials (RCTs) and observational studies data and optimize their informativeness.

**Methods:** We developed an extract-transform-load data integration pipeline to combine 13 source databases with 31,786 patients: the “mother” and “high-definition” cohorts from the French MS registry and eleven industrial RCTs. We aimed to inform each treatment class initiation with at least 500 patients with 2-year clinical and MRI follow-up. Our data integration strategy used every patient visit as a potential baseline time point to inform a specific neurologist’ query to the platform, thus tailoring the actual analysis cohort to each patient.

**Results:** The resulting PRIMUS database had 12,953 patients with at least one informative visit. It could inform 7/8 common treatment initiation scenarios with at least 500 patients (range: 485 for glatiramer acetate; 1,754 for natalizumab). The per-visit integration identified 696 more patients in the high-definition cohort than the classical epidemiological per-patient integration (+114 %). Although the mother cohort’s longitudinal data were deemed to be sparse, we identified 6,128 informative patients (yield: 27.8%; mean: 2.2 visits per patient).

**Interpretation:** A data integration pipeline and per-visit integration enabled us to build a highly informative reference database to be queried by neurologists through a web application to support discussions with their patients and the selection of disease-modifying treatments.

## Introduction

Chronic diseases are marked by heterogeneous activities, progressions, and therapeutic responses. In diseases with unsolved mechanisms, the data-driven approach of precision medicine aims to leverage panels of statistical biomarkers ^1^ and software providing clinicians with individual predictions, called clinical decision support systems (CDSS) ^2,3^. This requires the secondary use and combination of multiple sources of longitudinal individual patient data to achieve informative high-volume and high-quality reference databases ^4,5^. Data integration pipelines are needed to harmonize multi-source data into a common data model with scalable strategies ^6^ despite the various purposes of their primary data collections and prepare them consistently for a given analysis despite varying quality.

Compared to real-world data provided by top-down initiatives through health data warehouses (a.k.a., “hubs”) ^7^, research data are primarily collected in a structured fashion to produce scientific evidence, either through randomized clinical trials (RCT) or observational studies (OS). Patients may consent to secondary uses immediately from data collection. Industrial RCT data have the highest level of standardization. Their information content provides the highest level of evidence to approve disease-modifying treatments on the market ^8^. However, RCT volume is relatively modest, study populations are curated, typically testing only one or two active treatments, and longitudinal follow-up is short unless study extensions are carried out. OS data are actively collected in a bottom-up effort by research networks during routine practices with dedicated case report forms. These disease registries may collect data over the whole span of outpatient disease histories ^9^. They have varying quality but may reach high volumes and capture real-world practices including all the commonly prescribed treatments. Nested cohorts within the whole OS may have more standardized data collection.

The “Projections In Multiple Sclerosis” (PRIMUS) consortium is a partnership of academic and industrial stakeholders seeking to develop a precision medicine platform integrating OS and RCT data in multiple sclerosis (MS). It aims to make them accessible to neurologists during the consultation through a CDSS to support the selection of disease-modifying treatments based on the visualization of similar patient data ^10^. MS is the most frequent chronic autoimmune disease of the central nervous system with numerous approved therapeutic options ^11^. MS disease activity is characterized by relapsing episodes of acute symptoms and by the subclinical accumulation of demyelinating lesions in the brain and spinal cord assessed by magnetic resonance imaging (MRI). MRI data have become critical to monitor treatment response in routine practices with quantitative decision thresholds ^12,13^.

This study explored to what extent OSs of the French MS registry and industrial RCT data could be reused secondarily and combined to inform individual prognoses in MS through a CDSS. We describe the resulting database and the data integration pipeline we developed to achieve this integration in a scalable fashion and to optimize the information provided by the follow-up of each patient.

## Materials and Methods

### Source databases

We considered 13 source databases: two from the French MS registry (*Observatoire Français de la Sclérose en Plaques*; OFSEP; NCT03603457) ^14^; four RCTs from Merck ^15–18^; and seven RCTs from Biogen ^19–25^. The study designs and their population sizes are described in Figure 1 and their format in supplementary Figure 1. The French MS registry is a national-scale network of 40 neurology departments collecting data continuously since 2011 and a centralized coordination and data monitoring in Lyon ^26^. MRI data are collected from the routine radiologist’s naked eye readings. A subpopulation of 2,842 patients was included in a nested “High definition” cohort (OFSEP-HD) between 2018 and 2020 with a planned yearly follow-up of 5 years and standardized computerized MRI readings. Raw brain and spinal MRI images acquired with routine protocols were transferred from the local centers to the Shanoir platform based in Rennes and developed by the National Institute for Research in Digital Science and Technology (*Institut national de recherche en sciences et technologies du numérique*; INRIA) ^27^. Quantitative cross-sectional and longitudinal features about the T2 lesion load and activity were extracted from 11,656 brain MRIs with the Neuro.MS software (Pixyl SAS) ^28^. T1 and gadolinium-enhancing (GdE) lesions were not extracted. Another independent population of 22,011 patients selected from the main OFSEP “mother” cohort was also considered. This population aimed to explore the informativeness of an extended version of the PRIMUS database, whose data has been collected along routine practices in the OFSEP network and was therefore deemed to be sparse. At least 3 clinical visits and MRIs after January 1^st^, 2015 were required to include a patient in this source population. For patients in OFSEP-HD, the period from January 1^st^, 2015 to the inclusion in the nested cohort protocol has also been explored. Some raw MRI images could be transferred to the Shanoir platform on the initiative of the local centers but were not flagged a priori for standardized reading. The observation periods of the RCTs spanned from 2001 to 2015. Among the 9,617 patients of the RCTs intention to treat populations, we only integrated the 6,933 patients assigned to commercialized treatment regimens or placebo. We assessed 3 versions of the database: the unenriched OFSEP-HD cohort restricted to standard MRI readings (source number of patients: 2,842); the predefined version combining OFSEP-HD and the RCTs (source number of patients: 9,775); and the extended version allowing MRIs with naked-eye readings before and after the inclusion in OFSEP-HD and adding the OFSEP mother cohort (source number of patients: 31,786).

**Figure 1.**
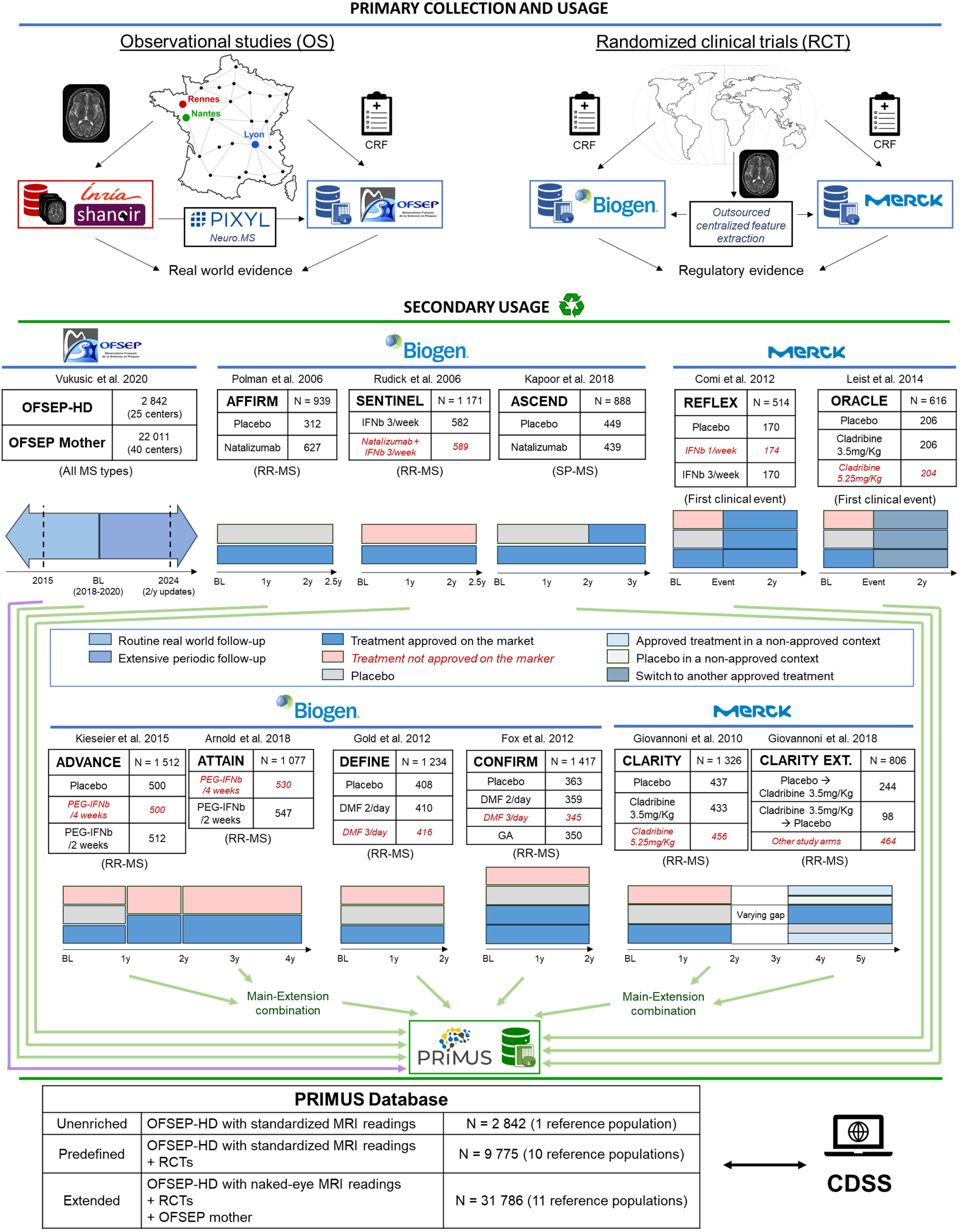
Data sources of the PRIMUS database and their study designs. Research data are primarily collected to generate evidence (Top). The PRIMUS project aims to develop the secondary usage of these data (Bottom) to be queried by neurologists through a clinical decision support system (CDSS). The 13 source studies are presented in bold with their intention to treat populations (i.e., the number of randomized and dosed patients). ATTAIN and CLARITY EXT were extension studies whose data have been combined with ADVANCE and CLARITY respectively, yielding 10 independent reference populations in the predefined PRIMUS database version (green arrows) and 11 in the extended version (green and purple arrows). Patients who received non-commercial treatment regimes were excluded (red). CRF: Case report form.

### Data integration pipeline

We developed ClinSci (Clinical Science), an extract-transform-load (ETL) pipeline to harmonize the source databases and integrate them into an analysis-ready format (Supplementary Figures 1-5). ETLs are software widely used in business intelligence data architectures to streamline database conversions between various data models ^29,30^: data lakes to store raw collected data, data warehouses to transform the data into an analysis-ready format, and data marts to expose data to be queried on-demand by decision-making users ^6^. Each source dataset is processed as a separate “batch” to make the integration process modular and to anticipate the distributed architecture of the PRIMUS data mart, where each data provider would host their partition. RCT extension studies were transferred as separate databases but were combined with the main study data at early steps. As a result, the end product of each batch is an independent reference population. For OFSEP-HD, both follow-up periods (before and after the nested protocol inclusion) were integrated into the same batch while differentiating both MRI readings.

### Per-visit integration

After harmonizing the data models of each batch, ClinSci enabled integrations into alternative analysis-ready formats. The technicalities are detailed in the supplementary materials. In the case of the PRIMUS CDSS, we compared per patient and per visit integration strategies (Fig 2). Per patient integration corresponds to the state-of-the-art practice of post hoc studies restricting the analysis to independent observations, taking only one time point as baseline for the analysis. However, MS is a chronic disease and one patient may go through several therapeutic scenarios during his/her follow-up. Per visit integration aims to leverage all meaningful visits by extracting several “segments of reference” (SORs) along a patient follow-up. SOR data are formatted like classical epidemiological studies: an index timepoint with cross-sectional baseline characteristics, a one-year past time window with longitudinal baseline characteristics, and two horizon timepoints at one and two years with cross-sectional and longitudinal endpoints. This corresponds to the classical durations of RCTs in MS (Fig 1). Two horizons were defined to control for the delay of action of some disease-modifying treatments (i.e., run-in period), as highlighted by ADVANCE ^23^ and epidemiological studies ^13^. The early disease period was enriched by a clinical visit, assuming no disability (Expanded Disability Status Scale; EDSS = 0) one year before the clinical onset, which serves as a past-time point in this context. Similarly to dynamic propensity score matching in epidemiological studies ^31,32^, each query of the CDSS to the data mart would build a personalized analysis cohort on the fly by selecting only the first SOR for each patient, if any matches the profile of the visiting patient (Fig 2).

**Figure 2.**
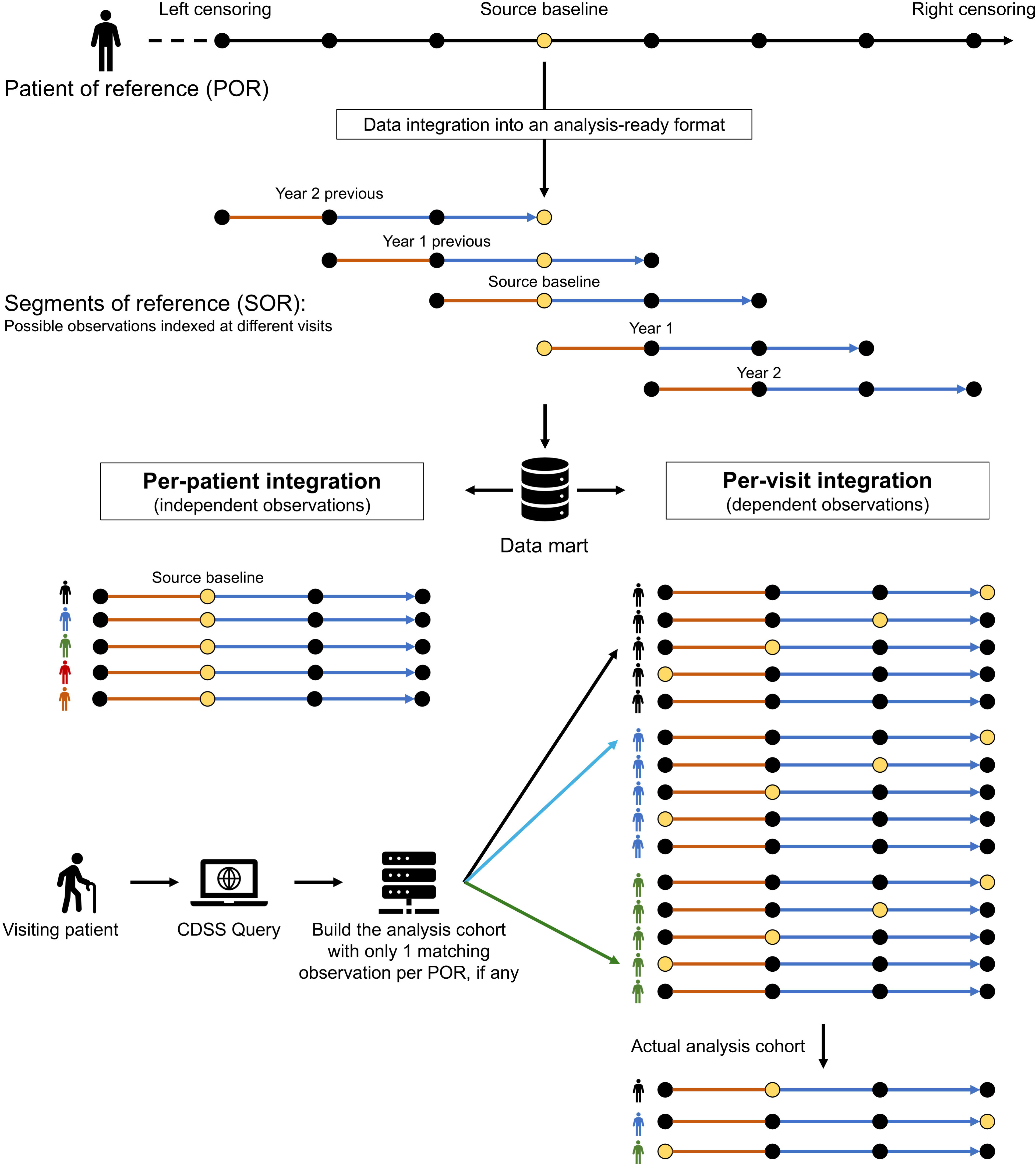
Per visit integration of the data mart by integrating the data as “segments of reference”. Several observation units can be extracted from the longitudinal data of one patient of reference (Top). Reference data have been formatted into multiple 3-year segments of follow-up per patient (i.e., segments of reference), each indexed from a different visit with a 1-year time window before the baseline visit and 2 years of subsequent follow-up. This yields a data mart with dependent observations that are not meant to be analyzed all at once by the CDSS (Bottom). With the per-visit integration strategy, each query of the CDSS to the datamart only selects the first segment of reference for each patient, if any matches the profile of the visiting patient, to dynamically build the actual analysis cohort at query time.

### Validation of the veracity of the integrated data

The veracity of the integrated data was assessed at the patient level through the visual inspection of individual timelines for consistency. For each reference population from an RCT, the reported efficacy endpoint analyses were replicated and checked for consistency with the publication results: relapse activity; T2 and GdE MRI activities; and confirmed disability worsening (CDW) as defined in the respective publications. We considered an analytical variability within the reported 95% CI and below an absolute error of 5 % to be satisfactory. The OFSEP populations did not have published references with the same transferred population. We assumed the veracity thanks to the consistency of the data transformations along the pipeline, the visual inspections of individual timelines, and continuous internal tests of a CDSS prototype by the authors with MS expertise ^33^.

### Informativeness assessment

We expressed the informativeness of the database as the number of observation units (i.e., patients or SORs) with complete data according to a predefined panel of predictors and outcome variables. The authors, who had clinical expertise in MS, elicited a predefined panel (Table 1). Our objective was to restrict the selection to variables identified as predictive in epidemiologic studies and collected in routine practice. The panel was then refined in a data-driven fashion based on the completeness of the data collection in each source (Table 1).

**Table 1.** Predefined and refined panels of predictors and outcome variables for the PRIMUS CDSS. The panel has been refined based on the inconsistencies of data collection across the different sources. In reference populations where only the aggregate of new and enlarging T2 lesions was collected, we neglected the number of enlarging lesions (IZ). The main refined panel included clinical and longitudinal MRI data, except the past disability evolution (all the variables not marked by an asterixis). Figure 4 explores a less stringent panel only with clinical data and 2 more stringent panels: one with the cross-sectional T2 lesion load (*) and one including the past disability evolution (**). The presence of spinal cord lesions has been left optional as it was only available in the OS sources (***).

Finally, we assessed the informativeness for each therapeutic scenario. Therapeutic scenarios were defined with a therapeutic class granularity as the conjunction of the treatment class received the year before the baseline time point and the treatment class received immediately after. In an intention-to-treat approach, the treatment class received after the baseline visit did not have to be maintained over the two following years. Scenarios with off-label treatments or several successive disease-modifying treatments during the previous year were considered ambiguous and have been excluded (5.2% and 8.2% of SORs regardless of the completion of the panel respectively in the predefined and extended databases). Our goal was to reach an informativeness of 500 unique patients for every clinically relevant scenario, which corresponds to the data volume of a phase 3 RCT arm.

### Ethical statement

The research was conducted under the consortium agreement of the ANR-21-RHUS-0014 PRIMUS project and the MR004 data processing regulation framework of the French Personal Data Regulatory Commission (Commission nationale de l’informatique et des libertés; CNIL).

## Results

### A scalable pipeline preserving the veracity of the information content

The ClinSci pipeline proved scalable by harmonizing two OSs and 11 RCT databases with a consistent data strategy, robust to the various study designs (OS against RCT; predefined follow-up duration against time-to-event). Although the Clinical Data Interchange Standards Consortium (CDISC) standards promote harmonized data modeling and variable naming conventions ^35^, its implementation in the raw dataset schemes was varying, yielding partial compatibility. The supplementary material describes the key processing steps of the pipeline (supplementary Fig 1 – 5).

We assessed the veracity of the information content of the resulting data mart by replicating the population-level analyses reported in the RCT publications (Fig 3). Almost all estimates were in the 95% CIs and there remained an absolute analytic variability of 4 % at worst. Only the proportion of CDW events in SENTINEL (beta-interferon arm and CONFIRM (dimethyl fumarate arm) exceeded the 95% CI. This may be due to the integration with discrete timing, the implementation margin of CDW events definitions, different statistics software, and the privacy-enhancing transformations upstream to the data transfer. We chose to prioritize the consistency of CDW events consolidation across all batches. Overall, these results were considered satisfactory. At the individual level, the integrated data were consistent with the source datasets at the inspection of 10 randomly selected individual timelines for each reference population.

**Figure 3.**
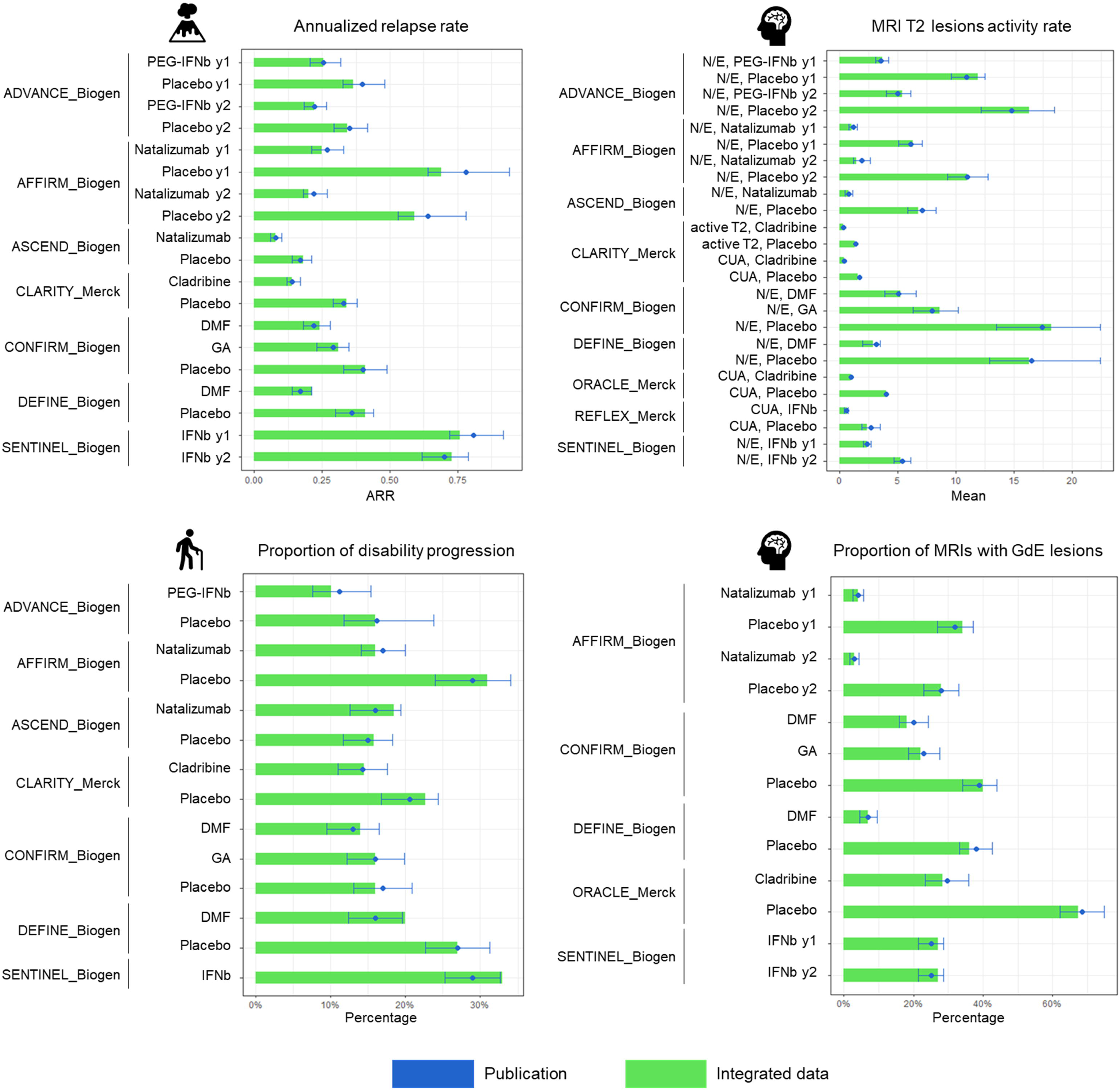
The veracity of the information content of the PRIMUS database. For each reference population, we formatted the batch data from the harmonized data warehouse into an analysis-ready format adapted to the replication of the reported population-level analysis of the endpoints of relapse activity, T2, and gadolinium-enhancing (GdE) activities, and confirmed disability worsening (CDW). All endpoints are over the 2-year follow-up period of the source study unless specified (e.g., y1 for endpoints at 1 year). The 95% confidence intervals are represented as error bars. If not explicitly reported in the publication, they were estimated from Gaussian laws. ARR: Annualized relapse rate; CUA: Combined unique active lesions; DMF: Dimethyl fumarate; GA: Glatiramer acetate; IFNb: Interferon beta; N/E: New or enlarging lesion; PEG-IFNb: Pegylated interferon beta.

### Per-visit integration increased the informativeness

For RCTs, the major limitation of informativeness regarding the predefined panel was the lack of MRI data and Expanded disability status scale (EDSS) rating for the year preceding the inclusion. Therefore refined panel used the GdE lesions count at the index visit as a proxy of the previous MRI activity and kept the past disability evolution as optional (Table 1). The cross-sectional T2 lesion count has also been considered optional as not collected in CONFIRM and ASCEND. Nevertheless, the informativeness of the predefined database remained much lower than the 9,775 source patients (Figure 4A). One factor is the 1-year and 2-year attrition (Fig 4B). The time-to-event trials REFLEX and ORACLE could have follow-ups shorter than 2 years, with respectively 14.0% and 54.9% 2-year attrition. There was from 7.8% to 22.8% attrition at 2 years in the other RCTs. Among the 2,842 OFSEP-HD patients, 2,677 (94.2%) patients had at least one MRI with standardized reading and 2,282 had one at the inclusion in the nested protocol (80.3%). Only a minority of them had an MRI with standardized reading the year before the inclusion in the nested protocol (n = 875; 30.8%).

**Figure 4.**
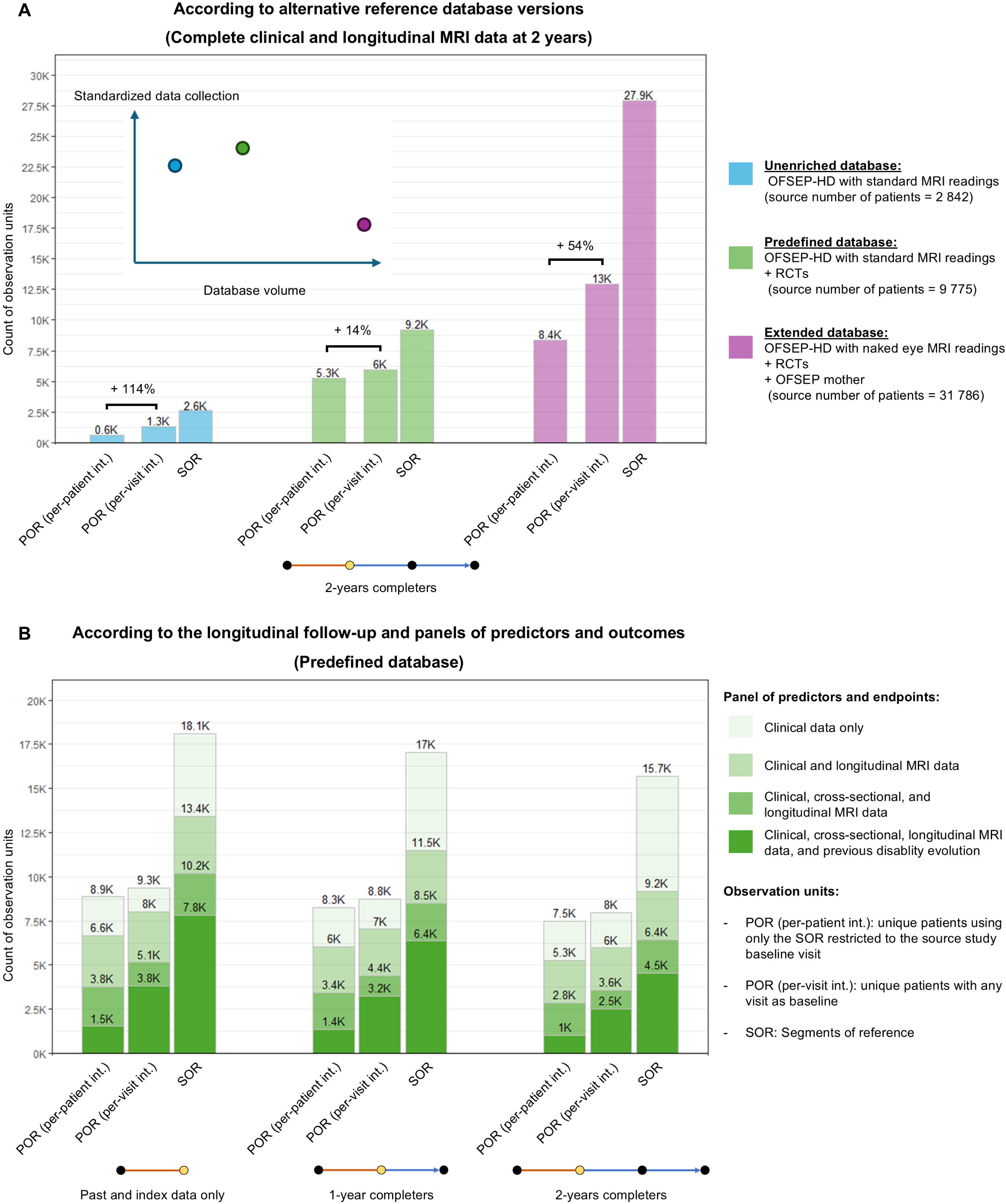
Per-visit integration increases the informativeness of the PRIMUS database. We assessed the informativeness as the number of observation units in different versions of the database (A). Per-visit integration enabled to identification of more patients with at least one complete SOR, especially in observational studies. Attrition and data incompleteness decreased the informativeness (B). The three major bottlenecks were the availability of MRI activity, MRI cross-sectional lesion load, and disability evolution data over the 3 years of a SOR. POR: Patient of reference; RCT: Randomized clinical trial; SOR: Segment of reference.

The 2020 COVID-19 pandemic has also disturbed the periodic yearly MRI follow-up. As such, the OFSEP-HD baseline yielded a SOR with a complete refined panel at 2 years for only 610/2.282 patients (26.7%). In the predefined database version, the enrichment by the 11 RCTs yielded 4,644 further patients with complete SORs against 6,933 patients in their cumulated intention-to-treat population (67.0%). Therefore predefined database had a total of 5,254 informative patients with per-patient integration restricted to the study baselines against 9,775 patients in the intention-to-treat population (53.7%).

We explored two mitigation strategies to engineer a more informative database. The first was per-visit integration. Enabling the use of any yearly visit as a potential baseline time point yielded complete SORs in 696 additional OFSEP-HD patients (+114 %; Fig 4A), showing that many series of successive yearly MRIs with standard readings are shifted from the predefined cohort baseline. Up to 5 SORs per patient could be extracted with a complete refined panel (mean = 1.5 SORs). RCT extensions of ADVANCE and CLARITY could also be leveraged. The second strategy was to increase the data volume with the extended version of the database. Allowing naked-eye MRI readings in OFSEP-HD before and after the inclusion in the nested protocol identified 861 additional patients with at least one complete SOR (+65.9%). Although the population of the OFSEP mother cohort was deemed to have sparse longitudinal data, 6,128 patients with complete SORs could be found with per-visit integration among the 22,011 patients in the source data (yield: 27.8 %; mean number of SORs per patient: 2.3; range: 1 - 9), against 2.218 patients with per-patient integration restricted to an arbitrary baseline set in 2019 to match the inclusion period of the OFSEP-HD cohort (yield: 10.1%). This yielded 12,953 unique patients in the extended database against 5,964 in the refined database (+ 117%; Fig 4).

### Uneven informativeness of the sources for the predefined panel

We further assessed the informativeness at the level of each variable related to the predefined and refined panels (Fig 5). Only subpopulations of CONFIRM and DEFINE had MRI follow-ups. Data collection in RCTs lacked spinal MRI, anteriority for MRI activity, and disability progression. The variables collected during the studies were also inconsistent, depending on the pre-defined analysis. The most potent example is T2 activity, which may be assessed as the number of new T2 lesions, the number of enlarging T2 lesions, or both aggregated. ADVANCE and ASCEND only collected the aggregate, requiring the refined panel to neglect the number of enlarging lesions in these studies. The cross-sectional T2 lesion load was also inconsistent. It was not collected in CONFIRM and ASCEND. In cases it was collected, it could have been assessed either in a quantitative fashion or just in a stratified fashion with a 9-lesions threshold in AFFIRM and SENTINEL. This shows how RCT data collections are focused on the pre-defined analysis.

**Figure 5.**
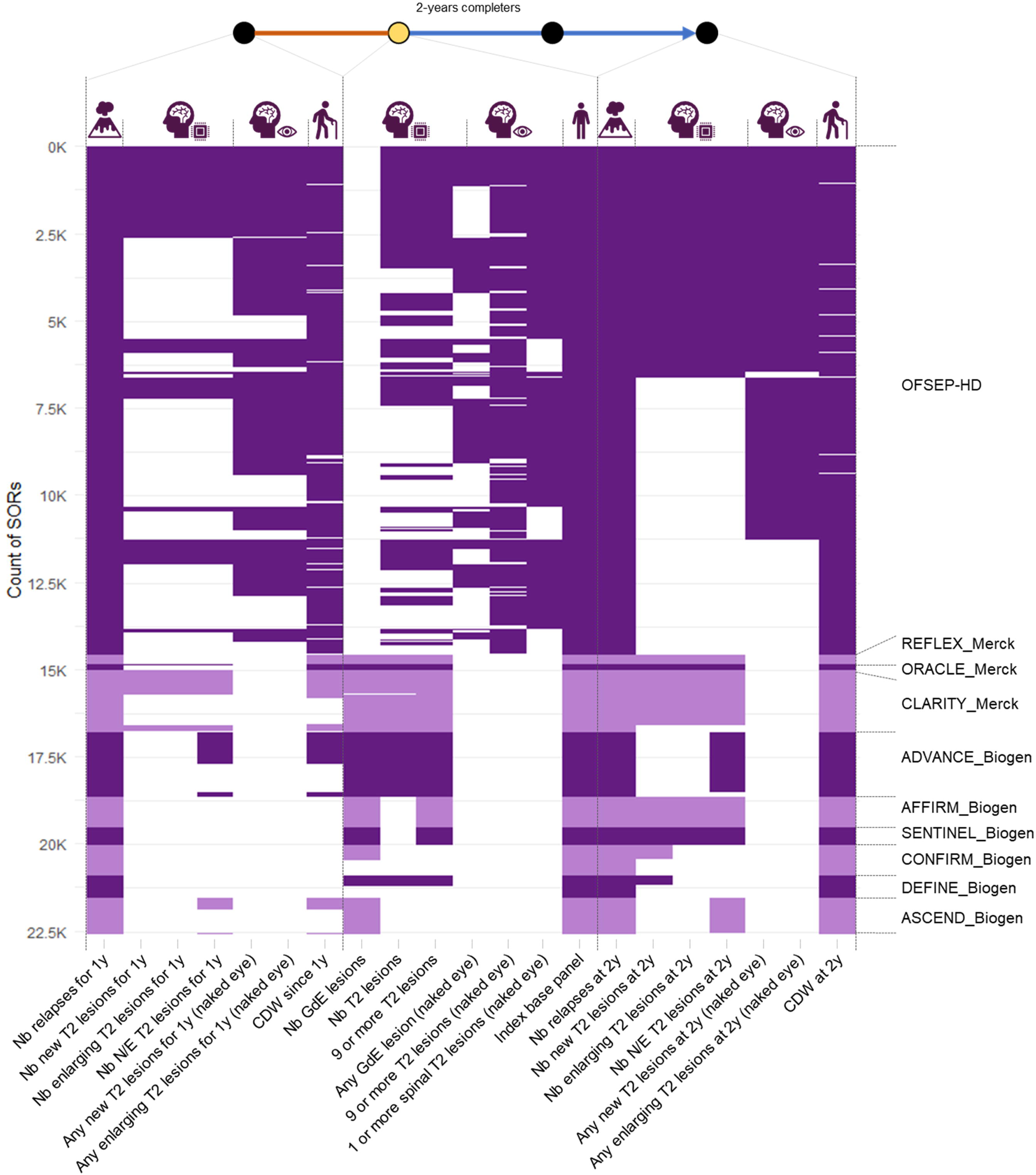
Uneven informativeness of the extended PRIMUS database. The missing data patterns (blank spaces) show the inconsistency of the data collection across and within the reference populations (alternative bands of color). Only the segments of reference (SORs) with complete relapse activity data are shown (n = 22,589). The OFSEP mother cohort is not represented for readability. Its missing data patterns are similar to OFSEP-HD with only naked-eye MRI readings. Only the variables about relapses, MRI activity (brain unless specified), and disability progression are detailed. The data at the 1-year horizon are not displayed for simplicity. The index base panel included sex, age at onset, disease duration, EDSS, pregnancy status, the past, and the index treatment. In OS and RCTs, some MRIs may have a stratified but not quantitative count of T2 lesions due to interpolations and extrapolations at data integration. CDW: Confirmed disability worsening; GdE: Gadolinium-enhancing; N/E: new or enlarging; RCT: randomized clinical trial.

Informativeness was also uneven when assessed for each therapeutic scenario (Fig 6). At their source baseline, RCTs informed only treatment starts as most studies included treatment-naïve patients or after a significant therapeutic window. Most OFSEP-HD patients maintained their treatments at the inclusion in the nested protocol. While the lack of informativeness for theoretical scenarios was expected, the low informativeness of the start of anti-CD20 and anti-anti-sphingosine-1-phosphate (S1P) drugs was considered to be a major issue for the PRIMUS project. Per-visit integration moderately increased the informativeness of some scenarios while the extended database version led to a major improvement. Even in the extended database with per-visit integration, only the treatment start scenarios enriched by RCT data reached the informativeness objective of 500 unique patients or more. Yet, high-efficacy treatments were historically positioned as second-line therapies, explaining why the informativeness is diluted in escalation switch scenarios. When aggregating the previous treatments and excluding maintenance scenarios in the extended database, the informativeness exceeded 500 unique patients for all common treatment classes except glatiramer acetate (range: 485 unique patients for glatiramer acetate; 1,754 unique patients for natalizumab). This range excluded rarely prescribed immune reconstitution therapies: alemtuzumab, mitoxantrone, and autologous stem cell transplantation. Some treatment discontinuation scenarios have also become informed by the extended database. They could be sustained in the long run or temporarily, such as long therapeutic windows, pregnancies, increased infusion intervals, or missing treatment data. Overall, the informativeness of the database varied greatly depending on the therapeutic scenario. The informativeness regarding the other patient characteristics has been described in Table 2.

**Figure 6.**
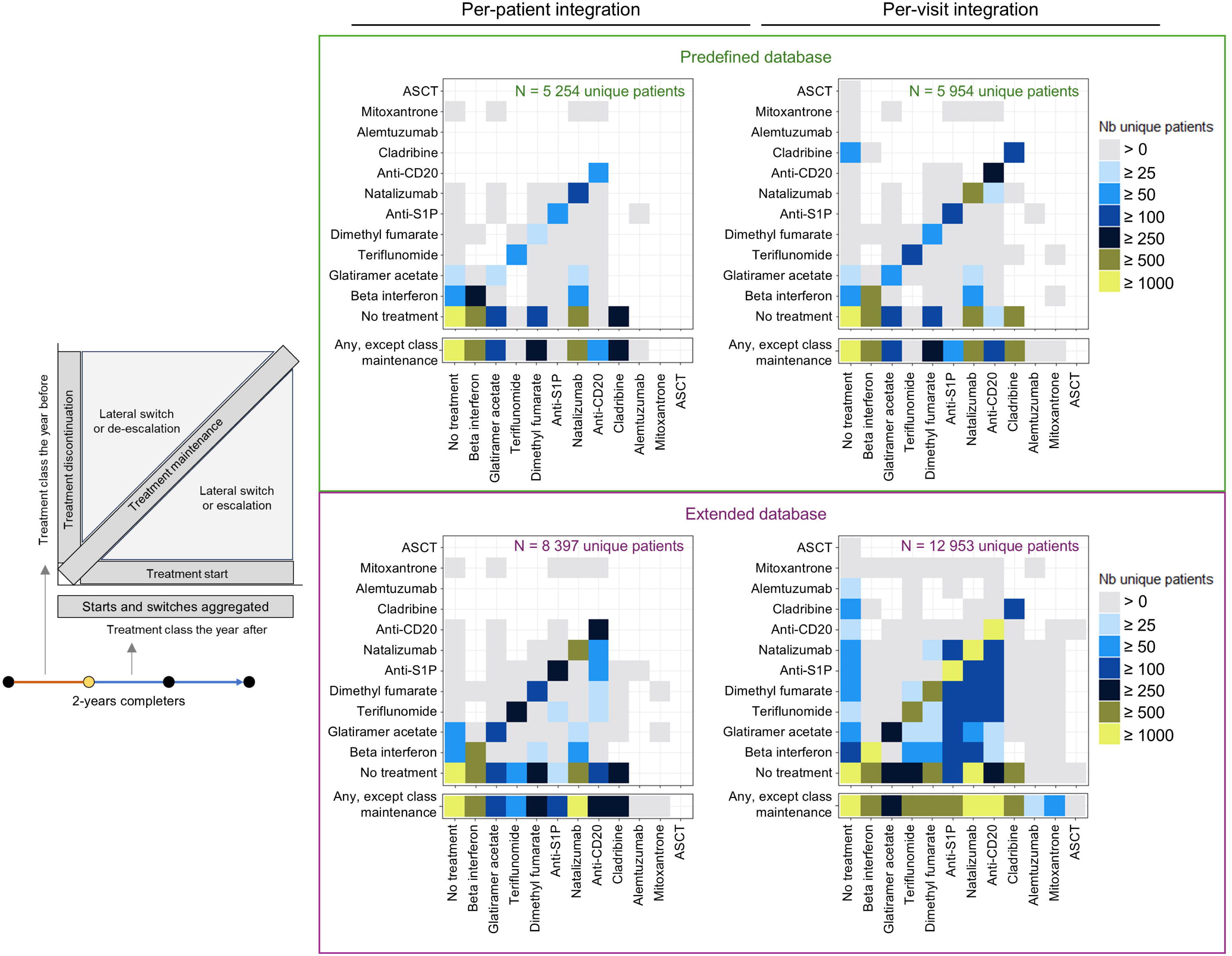
Informativeness of the PRIMUS database per therapeutic scenario. The therapeutic scenarios are defined with a therapeutic class granularity as the conjunction of the treatment received the year before the index timepoint and the treatment received after. All approved treatment regimes in MS are monotherapies. Scenarios with the succession of several active treatments during the previous year have been considered ambiguous and excluded. Placebo was assimilated to “No treatment”. Our goal was to reach at least 500 unique patients per clinically relevant scenario (yellow). ASCT: Autologous stem cells transplantation; S1P: anti-sphingosine-1-phosphate.

**Table 2.** Informativeness of the PRIMUS database per variable of the refined panel (except therapeutic scenarios). The results are restricted to segments of reference (SORs) with the complete refined panel. Due to the per-visit integration, the sum of unique patients over the strata of a variable may exceed the number of unique patients in the whole database. Therapeutic scenarios are described in Figure 6. EDSS: Expanded disability status scale; GdE: Gadolinium enhancement; PP-MS: Primary progressive multiple sclerosis; RIS: Radiologically isolated syndrome; RR-MS: Relapsing-remitting multiple sclerosis; SP-MS: Secondary progressive multiple sclerosis.

## Discussion

We explored the potential of secondary usage of 13 existing research databases in MS to create a Big Data resource capable of informing treatment selection through a CDSS. The resulting PRIMUS database can be assessed in the light of the 5 Vs of Big Data. (1) The enrichment of an OS database with RCT data yielded an important volume with 12,953 patients whose complete clinical and MRI activity data can inform 2-year individual prognoses in at least one context. (2) It integrated the variety provided by both clinical and MRI modalities of follow-up. (3) The veracity of the information content has been validated for RCTs and could reasonably be extrapolated to OS data thanks to the consistency of the data transformations with the ClinSci pipeline and the visual inspection of individual timelines. (4) The data mart is ready to be deployed in a cloud-based software architecture that will provide neurologists with high-velocity and continuous access to the data through the intermediate of the PRIMUS CDSS ^33^. (5) The informativeness assessment measured the value of the PRIMUS database to find similar patients across the whole spectrum of therapeutic scenarios. The real time analyses of these subgroups by the CDSS will be visual supports for shared decision making. It will also be a resource to develop other predictive analytics. The Evidence-Based Decision Support Tool in Multiple Sclerosis (EBDIMS) is another use case of integration of multiple studies into a reference database enabling neurologists to find similar patients with MS ^36^. However, it was restricted to clinical data and to placebo arms to inform the natural disease history. Other elements will contribute to the final value of the PRIMUS CDSS such as the validation of the predictive power of the analytics and the assessment of its clinical utility.

The ETL approach of the ClinSci pipeline streamlined our data integration framework. It went beyond the mere harmonization of databases to a common data model in medical informatics efforts to integrate the data for a clinical application of these databases. It brought consistency despite the diverse organizations, the incomplete application of industrial standards, and the study design specificities. We therefore have good confidence that ClinSci would be transferable to similar projects in a wide range of diseases. Knowing the high variability of data processing strategies ^37^, like in imaging ^38^, the inclusion of clinical domain expertise has been advocated as early as possible in clinical data pipelines to mitigate the risk of analytical variability and to fortify the correctness of the operations ^39^. While ClinSci’s framework gave traceability of the expert micro-judgments, it remained highly demanding in terms of technical skills and medical knowledge for the pipeline user. At this stage, it may serve as a code basis rather than software to ease the integration of multi-source research data into precision medicine platforms and thus promote their secondary uses.

Per-visit integration has been critical in extracting informative SORs from OS data by identifying complete 3-year series of clinical and MRI visits, especially in the OFSEP mother cohort, whose longitudinal data were deemed a priori to be too sparse to be informative for PRIMUS. The bottleneck of MRI data completeness highlights the need to streamline the collection of MRI data and feature extraction. Although the OFSEP mother cohort lacked standardized MRI readings, most raw MRI images are likely stored in the information system of each local hospital and a significant proportion may have already been transferred to the research neuroimaging Shanoir platform on the initiative of the local OFSEP centers. As a consequence of the results of this study, we have committed to a national data linkage effort to retrieve the raw images and expand the standardized MRI reading to the OFSEP mother cohort as much as possible. Our data integration pipeline with per-visit integration will be essential to screen the OFSEP registry and narrow this data linkage effort down to patients with promising SORs and the corresponding periods of follow-up.

Data collection is an active and costly process in medicine. It is therefore restricted to features with a priori known importance and accessible in routine, except in low-volume deep phenotyping cohorts such as the “very high definition” nested cohort of OFSEP (ongoing recruitment aiming at 250 patients). In RCTs, the inconsistent data collection can be explained by the different stages of MS. For instance, the cross-sectional T2 lesion load was especially collected in the RCTs interested in the early stages of MS because it was part of the Barkhof criteria to predict the risk of a second relapse ^40^, whereas the past disability evolution was only screened in the RCT including patients with SP-MS. This led in our case to an uneven informativeness for secondary usage. Even with approximations and proxies in the refined panel of variables, data were still highly incomplete. Complete datasets are required to develop model-based analytics because all SORs would be processed likewise. The solution we plan to investigate is a data visualization analytic that would personalize the usage of each SOR. Contrary to model-based analytics, clinical reasoning typically relies on different panels of predictors in different contexts of a clinical pathway ^2^. We reported a proof of concept of filter-based data visualization analytics with ADVANCE as the only reference population ^10^. It leaves the neurologist the choice of the analysis definition by activating each filter or not, based on what is considered clinically relevant. There is ongoing work by the authors to design a human-machine interface with elements to provide data-driven guidance to navigate the reference data.

Another challenge of multi-source data integration is the mitigation of population effects. The PRIMUS CDSS aims to provide indirect treatment comparison, which consists of inferring treatment effects by comparing the measures obtained from different reference populations. Network meta-analysis is a classical method when only population-level data are available ^41^, but it neglects numerous confusion factors that may arise from (1) the difference in characteristics of the populations and (2) the lack of head-to-head standardization of the data collection. Methods like matching-adjusted indirect comparison have been proposed in cases where individual patient data is available for one population ^42^. The indirect comparison would be restricted to a subgroup matching the population-level characteristics of the second population. The filter-based data visualization analytic of PRIMUS would provide a similar matching. We plan to investigate statistical recalibrations of the residual biases in a study dedicated to the analytic layer of the PRIMUS platform.

In the near future, we aim to improve the informativeness of PRIMUS by integrating as many phase 3 RCT databases as possible. In the long term, foreign OSs with standardized MRI readings could also be considered. The ClinSci pipeline would streamline updates of the data mart as new databases will be integrated from RCTs and OSs. With further software development, data integration could be decentralized to avoid data transfers by running ClinSci in environments hosted by the data owners. There is ongoing work by the PRIMUS consortium to collect RCT databases from “data sharing” platforms^43^. However, the “data sharing” of these platforms is restricted for regulatory reasons to punctual data usage in virtual working environments, like classical post hoc studies. Data visualization analytics require continuous access to individual patient data to support routine practices. Synthetic data is an emerging privacy-enhancing technology that could yield a fully anonymous data mart ^44,45^. This could overcome the regulatory sensitivity of individual patient data to unlock their sharing and value to inform precision medicine.

## Conclusion

We described the data integration aspect of the PRIMUS project as an effort to leverage the currently existing databases in MS to inform precision medicine. The resulting database will be a resource of high-granularity information to be queried in real-time by neurologists through a CDSS to support discussions with their patients and the selection of disease-modifying treatments. The high but uneven informativeness achieved questions the reality of a Big Data regime in the field of MS and chronic disease management in general. We reported a data integration pipeline and an integration strategy to get closer to such a regime, where physicians would support their decisions and discussions by navigating individual patient data in real-time through precision medicine platforms.

## Acknowledgements

We thank Biogen, Merck, and OFSEP for providing the source data. We also thank Nathalie Blanc, Sabrina Banon, and Joelle Martin-Gauthier for their support in project management. We are most grateful to the Genomics Core Facility GenoA, member of Biogenouest and France Genomique and to the Bioinformatics Core Facility BiRD, member of Biogenouest and Institut Français de Bioinformatique (IFB) for the use of their resources and their technical support.

This work is part of the PRIMUS project, which was supported in part by the French National Research Agency (Agence Nationale de la Recherche, ANR) as its 3rd PIA, integrated to France 2030 plan under reference [ANR-21-RHUS-0014].

The data collection of OFSEP has been supported by a grant provided by the French State and handled by the “Agence Nationale de la Recherche,” within the framework of the “France 2030” program, under the reference ANR-10-COHO-002”, and the support of the “Eugène Devic EDMUS” foundation against multiple sclerosis.

## Authors contributions

S. Demuth: Conceptualization, Data curation, Formal analysis, Investigation, Methodology, Writing – original draft.

IF: Data curation, Formal analysis, Investigation, Writing – review & editing.

JP: Writing – review & editing.

OR: Methodology, Validation.

NV: Writing – review & editing.

SL: Writing – review & editing.

RC: Resources, Methodology, Writing – review & editing

AK: Funding acquisition, Supervision, Validation, Writing – review & editing.

BB: Resources, Writing – review & editing

MP: Resources, Writing – review & editing

SV: Funding acquisition, Resources, Writing – review & editing.

S. Doyle: Funding acquisition, Resources, Software

GJ: Resources, Software

JDS: Supervision, Writing – review & editing.

DL: Funding acquisition, Supervision, Validation, Writing – review & editing.

GE: Funding acquisition, Supervision, Validation, Writing – review & editing.

PAG: Conceptualization, Funding acquisition, Methodology, Supervision, Validation, Writing – review & editing.

## Data availability

ClinSci has been made available at GitLab ^46^. For reference, the configuration files have been made available (except for the patient identifiers mappings). The databases could not be made publicly available.

## Abbreviations

CDSS: clinical decision support systems
CDW: confirmed disability worsening
CDISC: Clinical Data Interchange Standards Consortium
ClinSci: Clinical Science
EDSS: Expanded disability status scale
ETL: extract-transform-load
GdE: gadolinium-enhancing
MRI: magnetic resonance imaging
MS: multiple sclerosis
OFSEP: Observatoire Français de la Sclérose en Plaques
OS: observational studies
PRIMUS: Projections In Multiple Sclerosis
RCT: randomized clinical trials
SOR: segments of reference

