## Supplementary material for "Are we in a Big Data era for multiple sclerosis? Lessons from integrating clinical trials and observational studies data into the PRIMUS precision medicine platform": Table 1

| **Time point of the SOR** | **Predefined panel** | **Modifications for the refined panel** |
| --- | --- | --- |
| Past time window  (1 year) | Number of relapses |  |
|  | Number of new T2 lesions in the brain | Presence of new T2 lesions in the brain  Or  Presence of new or enlarging T2 lesions in the brain ✝  Or  Presence of GdE lesions in the index MRI |
|  | Disability evolution | ** |
|  | Disease-modifying treatment covering the time window |  |
| Baseline time point | Sex |  |
|  | Age at clinical onset |  |
|  | Disease duration |  |
|  | MS type |  |
|  | Number of T2 lesions in the brain | Number of T2 lesions in the brain stratified to 9 lesions * |
|  | Number of T2 lesions in the spinal cord | Presence of any spinal cord T2 lesion *** |
|  | Pregnancy status |  |
|  | EDSS |  |
|  | Ongoing disease-modifying treatment |  |
| Horizons  (1 and 2 years) | Number of relapses |  |
|  | Number of new T2 lesions in the brain | Presence of new T2 lesions in the brain  Or  Presence of new or enlarging T2 lesions in the brain ✝ |
|  | Disability evolution |  |
|  | Ongoing disease-modifying treatment | Ongoing disease-modifying treatment (for OS data only) |

**Table 1. Predefined and refined panels of predictors and outcome variables for the PRIMUS CDSS.** The panel has been refined based on the inconsistencies of data collection across the different sources. In reference populations where only the aggregate of new and enlarging T2 lesions was collected, we neglected the number of enlarging lesions (✝). The main refined panel included clinical and longitudinal MRI data, except the past disability evolution (all the variables not marked by an asterixis). Figure 4 explores a less stringent panel only with clinical data and 2 more stringent panels: one with the cross-sectional T2 lesion load (*) and one including the past disability evolution (**). The presence of spinal cord lesions has been left optional as it was only available in the OS sources (***).
