## Supplementary material for "Are we in a Big Data era for multiple sclerosis? Lessons from integrating clinical trials and observational studies data into the PRIMUS precision medicine platform": Table 2

|  |  | **Predefined database** | | **Extended database** | |
| --- | --- | --- | --- | --- | --- |
| **Time point of the SOR** | **Variable** | **SORs**  **(n = 9 185)** | **Unique patients**  **(n = 5 954)** | **SORs**  **(n = 27 916)** | **Unique patients**  **(n = 12 953)** |
| Past time window  (1 year) | Number of relapses |  |  |  |  |
|  | 0 | 4 624 | 2 639 | 18 996 | 8 523 |
|  | 1 | 3 177 | 3 007 | 6 724 | 5 885 |
|  | >= 2 | 1 384 | 1 363 | 2 196 | 2 110 |
|  | MRI activity (T2 or GdE) |  |  |  |  |
|  | No | 5 750 | 4 118 | 19 673 | 10 039 |
|  | Yes | 3 435 | 2 664 | 8 243 | 5 930 |
|  | Disability evolution |  |  |  |  |
|  | Stable or improving | 4 385 | 2 515 | 20 249 | 8 683 |
|  | Worsening | 499 | 491 | 1 750 | 1 607 |
|  | Missing | 4 301 | 4 272 | 5 917 | 5 879 |
| Baseline time point | Sex |  |  |  |  |
|  | Female | 6 424 | 4 174 | 20 305 | 9 282 |
|  | Male | 2 761 | 1 790 | 7 611 | 3 671 |
|  | Age at clinical onset |  |  |  |  |
|  | < 20y | 1 080 | 707 | 3 613 | 1 623 |
|  | 20 - 40y | 6 558 | 4 311 | 19 457 | 9069 |
|  | >= 40y | 1 547 | 946 | 4 846 | 2 261 |
|  | Disease duration |  |  |  |  |
|  | < 2y | 1 266 | 1 119 | 4 216 | 3 060 |
|  | 2 - 10y | 4 318 | 2 850 | 12 011 | 6 275 |
|  | >= 10y | 3 601 | 2 344 | 11 689 | 5 457 |
|  | MS type |  |  |  |  |
|  | RIS | 1 | 1 | 220 | 215 |
|  | RR-MS | 7 847 | 5 144 | 24 350 | 11 396 |
|  | SP-MS | 1 242 | 811 | 2 696 | 1 464 |
|  | PP-MS | 95 | 55 | 650 | 303 |
|  | EDSS |  |  |  |  |
|  | <= 2.5 | 5 419 | 3 667 | 17 461 | 8 437 |
|  | 3 - 5.5 | 2 905 | 1 994 | 7 894 | 4 115 |
|  | >= 6 | 861 | 561 | 2 561 | 1 391 |
|  | Number of T2 lesions in the brain |  |  |  |  |
|  | < 9 | 506 | 382 | 2194 | 1 372 |
|  | >= 9 | 7 286 | 4 547 | 23 110 | 10 402 |
|  | Missing | 1 393 | 1 062 | 2 612 | 1 844 |
|  | Number of T2 lesions in the spinal cord |  |  |  |  |
|  | 0 | 290 | 162 | 2 850 | 1 546 |
|  | >= 1 | 1 960 | 1 002 | 14 714 | 5 947 |
|  | Missing | 6 935 | 4 854 | 10 352 | 6 473 |
|  | Pregnancy status |  |  |  |  |
|  | Not pregnant | 9 169 | 5 957 | 27 728 | 12 922 |
|  | Pregnant or pre-partum | 9 | 9 | 109 | 106 |
|  | Post-partum | 7 | 7 | 79 | 79 |

**Table 2. Informativeness of the PRIMUS database per variable of the refined panel (except therapeutic scenarios).** The results are restricted to segments of reference (SORs) with the complete refined panel. Due to the per-visit integration, the sum of unique patients over the strata of a variable may exceed the number of unique patients in the whole database. Therapeutic scenarios are described in Figure 6. EDSS: Expanded disability status scale; GdE: Gadolinium enhancement; PP-MS: Primary progressive multiple sclerosis; RIS: Radiologically isolated syndrome; RR-MS: Relapsing-remitting multiple sclerosis; SP-MS: Secondary progressive multiple sclerosis.
