## Supplementary for "Are we in a Big Data era for multiple sclerosis? Lessons from integrating clinical trials and observational studies data into the PRIMUS precision medicine platform"

### Supplementary materials

#### Source data

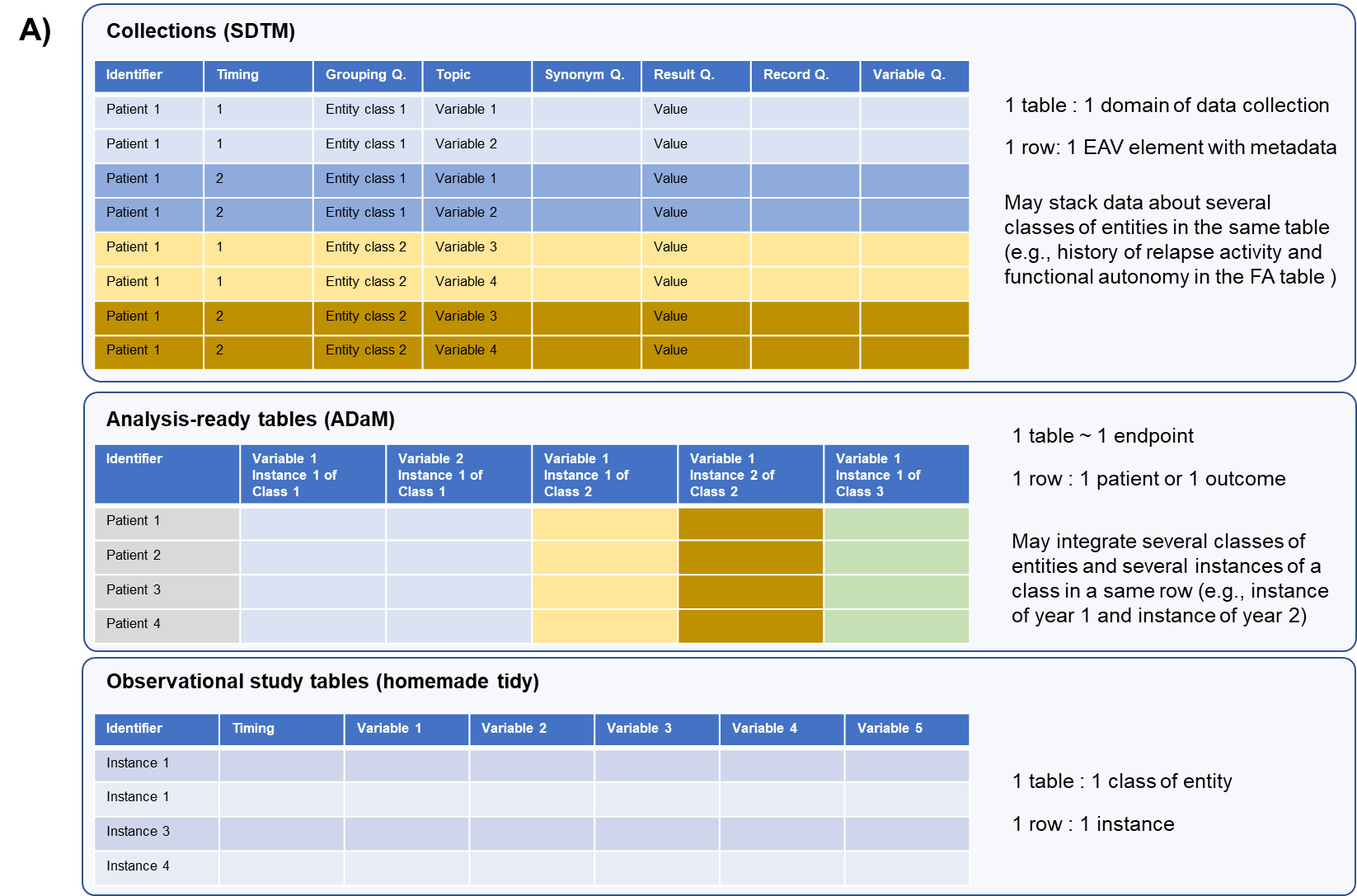

**Supplementary Figure 1.** The data from the 11 RCTs were transferred with standard CDISC data models: Study Data Tabulation Model (SDTM) and Analysis Data Model (ADaM). They respectively model data as collections of “findings” elements (colors represent observations for a given patient at a given visit) and analysis-ready tables. Data from the observational studies were transferred with homemade tidy data models. “FA” is a standard SDTM table name. CDISC: Clinical Data Interchange Standards Consortium; EAV element: Entity-Attribute-Value element; RCT: Randomized Clinical Trial.

#### The ClinSci pipeline

We implemented ClinSci as a sequence of Python scripts, each one performing a dedicated ETL transformation step (Supplementary Figure 2, Supplementary Table 1). Comma-separated values (CSV) configuration files enable the pipeline user to tailor the ETL transformations according to the batch specificities. The resulting intermediate versions of the database are stored as SQLite files. Additional transformation functions or algorithms, which may be written by the user, can be loaded as modules to perform feature engineering or rule-based consolidation. It also comprises the individual timeline viewer. For new batches, the configuration files and the intermediary database versions are generated gradually as the batch is being processed.

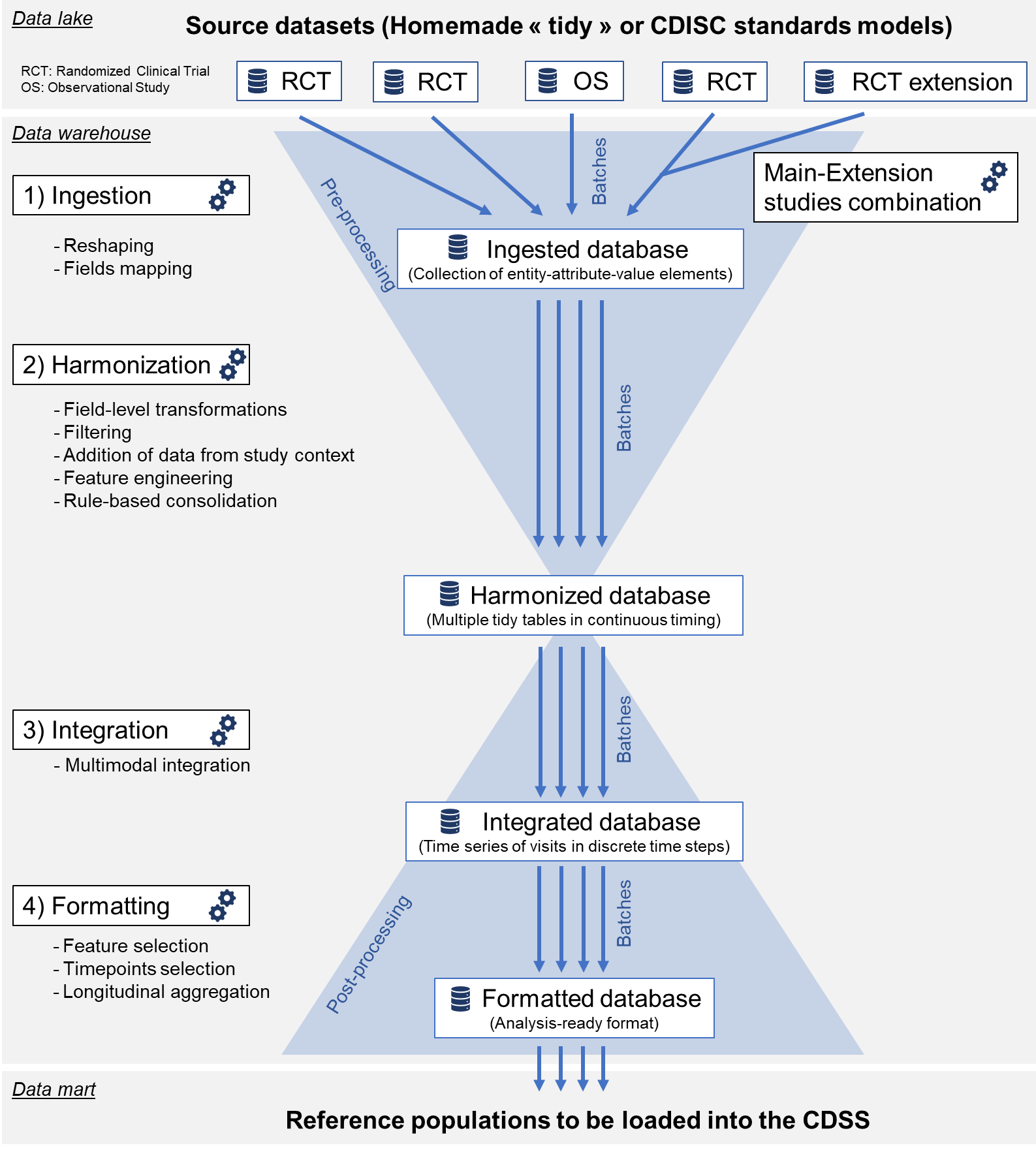

**Supplementary Figure 2. The ClinSci pipeline general structure.** Each source database is stored as received in a data lake and is then processed as a separated batch. The processing of a batch is a multistep process yielding intermediate versions of databases within a data warehouse. Data preprocessing (convergent part) focuses on data ingestion, harmonization, and cleansing. Data post-processing (divergent part) focuses on clinically meaningful data consolidation, integration rules, and feature selection. The post-processing is conceived as divergent because data become formatted (and thus aggregated) for a specific usage in a data mart. Alternative data usage (e.g., multiple CDSS analytics) may require different aggregations of data (e.g., analysis by the CDSS or the replication of RCTs analyses). Furthermore, following evolutions of expert consensus, several iterations may be enacted for a given usage. RCT: Randomized Clinical Trial; OS: Observational study, CDSS: Clinical Decision Support System.

#### Harmonization under a common data model

Data were ingested as collections of entity-attribute-value (EAV) elements (e.g., patient ID – birthdate – 1987-10-26) to perform table, field, and concept mappings (Suppleentary Figure 3). The harmonized common data model (CDM) of PRIMUS is a reshaped version into tidy tables according to relevant units of observations (i.e., one table per class of entity; one row per instance; one column per variable). The harmonization step comprised transformations to harmonize the content of the databases, such as data cleansing at the field level, and transformations to increase the compatibility across the batches by engineering inconsistently collected features.

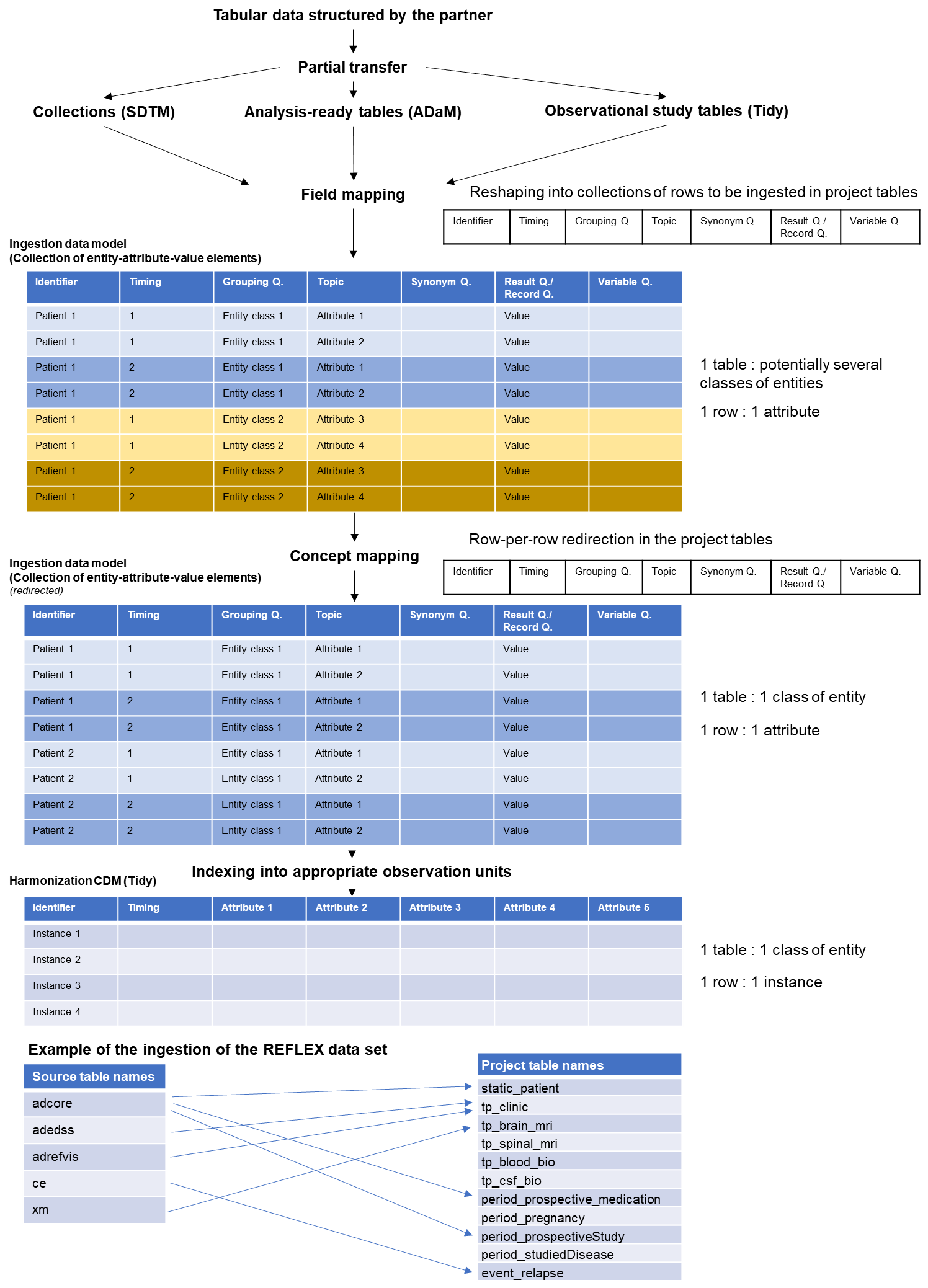

**Supplementary Figure 3.** Data ingestion process. Source tables were reshaped to ingest data as collections of entity-attribute-value elements (colors represent observations for a given patient at a given visit). This ingestion data model could include several levels of indexing and metadata. For SDTM tables, identifiers, timing variable, and grouping qualifiers were used as columns holding the entity indexes. Topic variables were used as columns holding the attributes. Synonym qualifiers hold verbose self-explanatory versions of the attribute names. Result and record qualifiers were used as the columns holding the values. Variable qualifiers hold metadata. The pipeline configuration requires the user to label ADaM and OS tables with these SDTM variable roles. This way, the pipeline splits entities of different classes that could have been concatenated in a column-wise fashion on the same row of ADaM or OS tables. It also splits ADaM rows when they combine data of several instances of an entity class (e.g., the experimental conditions of year 1 and 2). The entity-attribute-value elements were mapped to data warehouse tables defined by the platform project (one table per class of entity). We describe the example of the batch of the REFLEX trial at the bottom. The data warehouse table naming convention included the type of entity as prefix. “Static” entities are immutable attributes over the patient’s life, like patient demographics, or genotypes. “Tp” (for “Timepoints”) are time series of dynamic attributes collected at planned visits from various modalities of investigations, like clinical examinations, MRI scans, or laboratory tests. “Events” are punctual phenomena occurring outside planned visits, like MS relapses or adverse events. “Periods” span over a significant amount of time, like a treatment, a disease, or a study follow-up. They are characterized by a starting and ending date and may be decomposed into phases. Q.: Qualifier; CDM: Common Data Model. SDTM: Study Data Tabulation Model; ADaM: Analysis Data Model; OS: Observational study.

#### Consolidation of the harmonized data

MS experts of the PRIMUS consortium validated the rule-based consolidation algorithms (supplementary material; section MS-specific expert-defined criteria) during repeated interviews and focus groups. For instance, punctual treatment exposures were transformed into continuous periods of treatment (Supplementary Figure 4). In OSs, we used a modified version of the Lorscheider definition to compute the MS type at each clinical visit from the history of relapses and the EDSS trajectory ^1^.

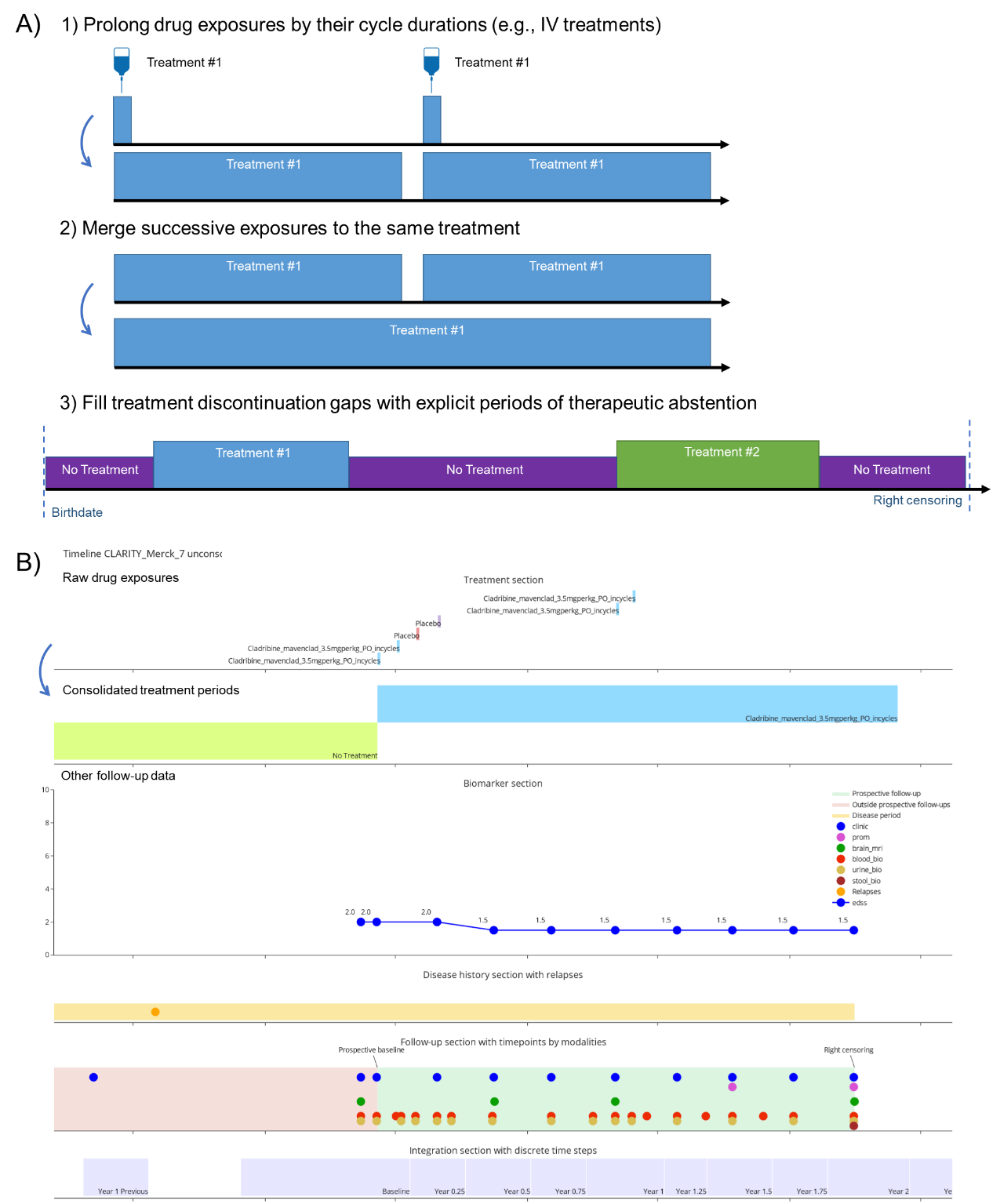

#### Supplementary Figure 4. Treatment consolidation. (A) General steps of the treatment consolidation algorithm. (B) Illustration of the algorithm result on a CLARITY patient individual timeline. In this dataset, the treatment “cladribine” and placebo exposures were recorded as doses. The algorithm merges the successive doses of a same treatment into continuous periods of treatment. It also prolongs the period after the last dose to account for the treatment activity over the administration cycles (for cladribine, it was set to 1 year). Some treatments (e.g., cladribine) may have a prolonged pharmacological effect, even after the end of the last administration cycle (i.e., induction effect). In this case, another phase may be described to account for the delayed wash-out of the treatment effect.

#### Data integration

The integration aggregated the data collected through different modalities of investigation into a single table representing a time series of “points of care” (Supplementary Figure 5). These “points of care” serve as time steps to transition from continuous to discrete timing. RCT visits were sufficiently periodic to label them according to time steps defined at the population level. In OSs, the time steps had to be defined at the patient level with a walking algorithm seeking the most clinically meaningful timepoint with 12-month intervals ± 6 months. Additional time points were created at each treatment change and considered to be the most meaningful. Events were integrated as cumulated counts since the inclusion of the patient in the study (source study baseline). Treatment periods are integrated according to their current phase at each time point (i.e., as joined lists of started, maintained, or stopped treatments during the time step associated with a time point; Supplementary Figure 5). To format the data into analysis-ready SORs, this “points of care” time series was reshaped. Cross-sectional variables were included as is, whereas the longitudinal variables are rebaselined relative to the index visit. Only yearly points of care were allowed as indexes with at least 2 years of expected follow-up according to the study protocol for RCTs or the date of data extraction for OSs.

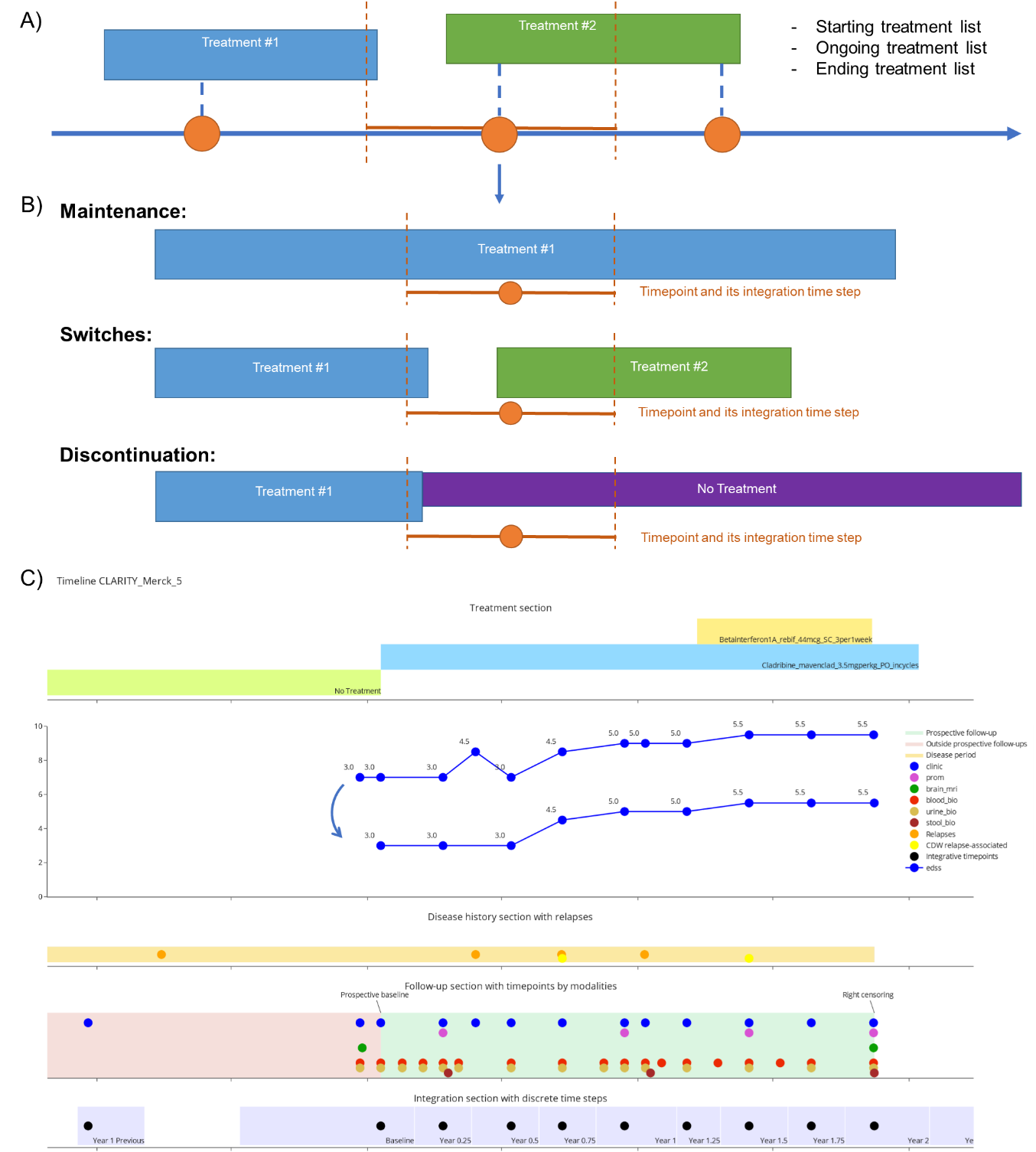

**Supplementary Figure 5.** Data integration by discrete time steps. (A) Timepoint, events, and period data are integrated over discrete time steps of follow-up defined by the pipeline user and centered by the expected timing of a visit (3-months time steps for RCTs) or defined automatically with a walking algorithm (personalized yearly time steps for OSs). (B) Illustration of the treatment data integration and the resulting interpretation of the therapeutic strategy. (C) Individual timeline of a CLARITY patient illustrating the integration of EDSS, the main disability scale in MS (higher values being worse). There could have been several measurements of the same variable during a 3-months time step. The EDSS trajectory is smoothened and relapse-associated reversible EDSS worsening are filtered out by the algorithm as we configured it to kept the lowest value measured during each time step. EDSS: Expanded Disability Status Scale. RCT: Randomized Clinical Trial.

#### MS-specific expert-defined criteria

##### Cycle duration of treatments:

The cycle duration of each treatment has been elicited with MS experts (supplementary table 2). Cladribine is an oral treatment but processed as an IV treatment because of an induction effect.

| **Treatment** | **Significant Duration (days)** | **Merge Proximity (days)** | **Cycle Length (days)** | **Induction Effect Duration (days)** |
| --- | --- | --- | --- | --- |
| Placebo | 0 | 30 | 0 | 0 |
| No Treatment | 0 | 30 | 0 | 0 |
| Betainterferon1A_rebif_44mcg_SC_3per1week | 30 | 90 | 0 | 0 |
| Betainterferon1B_betaferon_250mcg_SC_1per2day | 30 | 90 | 0 | 0 |
| Pegbetainterferon1A_plegridy_125mcg_SC_1per2week | 30 | 90 | 0 | 0 |
| Glatirameracetate_copaxone_40mg_SC_3per1week | 30 | 90 | 0 | 0 |
| Methotrexate | 30 | 90 | 0 | 0 |
| Azathioprine_imurel_100mg_PO_1per1day | 30 | 90 | 0 | 0 |
| Mycophenolatemofetil_cellcept_1g_PO_2per1day | 30 | 90 | 0 | 0 |
| Teriflunomide_aubagio_14mg_PO_1per1day | 30 | 90 | 0 | 0 |
| Dimethylfumarate_tecfidera_240mg_PO_2per1day | 30 | 90 | 0 | 0 |
| Diroximelfumarate_vumerity_462mg_PO_2per1day | 30 | 90 | 0 | 0 |
| Tolebrutinib_tolebrutinib_60mg_PO_1per1day | 30 | 90 | 0 | 0 |
| Fingolimod_gilenya_0.5mg_PO_1per1day | 30 | 90 | 0 | 0 |
| Ponesimod_ponvory_20mg_PO_1per1day | 30 | 90 | 0 | 0 |
| Laquinimod_laquinimod_0.6mg_PO_1per1day | 30 | 90 | 0 | 0 |
| Siponimod_mayzent_2mg_PO_1per1day | 30 | 90 | 0 | 0 |
| Cyclophosphamide_endoxan_750mgperm2_IV_1per5week | 0 | 90 | 35 | 0 |
| Natalizumab_tysabri_300mg_IV_1per4week | 0 | 90 | 30 | 0 |
| Rituximab_truxima_1000mg_IV_1per6month | 0 | 60 | 183 | 0 |
| Ocrelizumab_ocrevus_600mg_IV_1per6month | 0 | 60 | 183 | 0 |
| Ofatumumab_kesimpta_20mg_SC_1per4week | 0 | 90 | 30 | 0 |
| Tocilizumab_roactemra_8mgperkg_IV_1per1month | 0 | 90 | 30 | 0 |
| Cladribine_mavenclad_3.5mgperkg_PO_incycles | 0 | 183 | 365 | 4000 |
| Mitoxantrone_mitoxantrone_12mgperm2_IV_1per2months | 0 | 90 | 60 | 4000 |
| Alemtuzumab_lemtrada_12mg_IV_1per1day_5day | 0 | 90 | 365 | 4000 |
| AutologousStemCellTransplantation | 0 | 90 | 365 | 4000 |

**Supplementary Table 1.** Rule-based labeling of the treatments present in the datasets. Exposures shorter than the significant duration were excluded. Two sequential exposures of the same treatments were merged if the gap between them had no other active treatment and was shorter than the “Merge proximity”. We defined a “cycle length” for treatments administered periodically instead of daily. This enabled us to correct the exposure end date by extending it according to the cycle length. Some treatments are positioned as induction treatments (or immune reconstitution therapies), meaning that their pharmacological effect is expected to last longer than the end of the last administration cycle. We have added an induction effect with an arbitrary duration of several years to be able to account for distant exposures to these treatments.

##### MS stage transitions:

No consensus exist to define the date of the transition from relapsing-remitting MS (RR-MS) to secondary progressive MS (SP-MS). The MS stage at baseline was usually added from the study inclusion criteria. Data about the oligoclonal bands (OCB) status was missing in all RCT datasets. We assumed that all patients were OCB positive (1) in the light of the McDonald 2017 diagnostic criteria ^2^, using the OCB status as a proxy of the dissemination in time, (2) knowing the high prevalence of OCBs in MS patients (95%) ^3^, and (3) the primary context of usage of PRIMUS being clinical decision support for treatment selection rather than computer-aided diagnosis. As a consequence, we neglected the clinically isolated syndrome (CIS) stage, even in the RCTs recruiting patients after their first relapse to consider all patients who experience one relapse as relapsing-remitting MS (RR-MS). We used a modified version of the Lorscheider criteria to define the transition between RR-MS and secondary progressive MS (SP-MS) ^1^. Pyramidal functional system scores were not available in all batches to apply the original Lorscheider criteria. We interpreted disability worsening as punctual events of confirmed disability worsening (CDW). CDW was defined as a significant EDSS worsening (supplementary Figure 2): 1.5 points if the index EDSS was 0; 1 point if the it was between 1 and 5.5; 0.5 point if it was 6 or higher. To be confirmed, the EDSS value had to remain above this threshold for at least 6-months. In observational studies data, 6-months or more confirmation, rather than 3 months, showed more relevant results at individual timelines inspection. In the end, we defined the transition to SP-MS as the first 6-months CDW event independent of relapse activity above an EDSS 4. Patients with primary-progressive MS (PP-MS) were labelled so at OS dataset baseline. No RCT dataset investigated PP-MS.

##### T2 lesion activity:

There were inconsistencies in MRI data collection across the batches, especially for the assessment of T2 lesions activity. Studies could record the longitudinal features of T2 lesion activity either in two separated variables (new T2 lesions and growing T2 lesions), or aggregated in one, even in the SDTM table. To overcome this discrepancy, we adjudicated to neglect growing T2 lesions in the batches where both counts were aggregated.

##### T2 lesion load:

In RCTs, the cross-sectional count of T2 lesions was only collected at the baseline MRI, except in RCTs where the completion of McDonald criteria was a time-to-event endpoint. We therefore made an approximation by considering that new T2 lesions do not disappear, which enabled us to compute follow-up cross-sectional T2 lesions count by adding the longitudinal count of new T2 lesions to the baseline cross-sectional count. This shows how RCT data collections are focused on the pre-defined analysis.
